# Correlates of COVID-19 Vaccine Hesitancy among People who Inject Drugs in the San Diego-Tijuana Border Region

**DOI:** 10.1101/2021.10.29.21265669

**Authors:** Steffanie A. Strathdee, Daniela Abramovitz, Alicia Harvey-Vera, Carlos Vera, Gudelia Rangel, Irina Artamonova, Thomas L. Patterson, Rylie Mitchell, Angela R. Bazzi

## Abstract

**Background:** People who inject drugs (PWID) are vulnerable to acquiring SARS-CoV-2 but their barriers to COVID-19 vaccination are under-studied. We examined correlates of COVID-19 vaccine hesitancy among PWID in the U.S.-Mexico border region, of whom only 7.6% had received ≥one COVID-19 vaccine dose by September, 2021.

**Methods:** Between October, 2020 and September, 2021, participants aged ≥18 years from San Diego, California, USA and Tijuana, Baja California, Mexico who injected drugs within the last month completed surveys and SARS-CoV-2, HIV, and HCV serologic testing. Logistic regressions with robust standard error estimation via generalized estimating equations identified factors associated with COVID-19 vaccine hesitancy, defined as being unsure or unwilling to receive COVID-19 vaccines.

**Results:** Of 393 participants, 127 (32.3%) were vaccine hesitant. Older participants, those with greater food insecurity, and those with greater concern about acquiring SARS-CoV-2 were more willing to be vaccinated. Higher numbers of chronic health conditions, having access to a smart phone or computer, and citing social media as one’s most important source of COVID-19 information were independently associated with vaccine hesitancy. COVID-19-related disinformation was independently associated with vaccine hesitancy (adjusted odds ratio: 1.51 per additional conspiracy theory endorsed; 95% confidence interval: 1.31-1.74).

**Conclusions:** Nearly one third of PWID in the San Diego-Tijuana border region reported COVID-19 vaccine hesitancy, which was significantly influenced by exposure to disinformation. Interventions that improve accurate knowledge and trust in COVID-19 vaccines are needed to increase vaccination in this vulnerable population.

**Summary:** Nearly one third of people who inject drugs in the U.S.-Mexico border region reported COVID-19 vaccine hesitancy, which was significantly influenced by exposure to disinformation and social media and inversely associated with food insecurity and high perceived threat of COVID-19.

## Introduction

As the second year of the COVID-19 pandemic begins, an ongoing contributor to SARS-CoV-2 transmission is low vaccine uptake [1]. In high-income countries, structural issues relating to vaccine access, such as limited transportation and healthcare access, disproportionately affect under-represented minorities and those with low socio-economic status [2]. However, COVID-19 vaccine hesitancy has also emerged as a major problem, due at least in part to an “infodemic” of misinformation and disinformation [3]. Misinformation refers to inaccurate information shared without malicious intent, whereas disinformation is the deliberate spread of false information, for example, through organized social media campaigns [4]. Although both misinformation and disinformation have sowed confusion about the epidemiology of SARS-CoV-2 and severity of COVID-19 illness [3], anti-vaccine disinformation campaigns have seriously undermined public confidence in COVID-19 vaccine safety in the U.S. and elsewhere [5, 6], especially among Blacks and Hispanics [7].

People who use drugs, especially people who inject drugs (PWID), may be especially vulnerable to SARS-CoV-2 infection due to individual behaviors (e.g., using drugs with others, engaging in sex work [8]), elevated prevalence of chronic diseases [9], and homelessness, incarceration [8, 10], and other social and structural factors that limit healthcare engagement such as addiction-related stigma [11]. We previously reported that while over one third of PWID in San Diego County and Tijuana, Mexico had been infected with SARS-CoV-2, only 9% had received at least one COVID-19 vaccine dose [8].

To inform public health approaches to improve vaccine uptake, we studied COVID-19 vaccine hesitancy among PWID in the U.S.-Mexico border region. We hypothesized that socio-structural factors such as homelessness and Latinx ethnicity would be significantly associated with COVID-19 vaccine hesitancy, as well as COVID-19 misinformation. We were also interested in studying whether exposure to COVID-19 disinformation was significantly associated with COVID-19 vaccine hesitancy in a population that has historically had limited access to social media [12].

## Methods

### Participants and Eligibility

Between October 28, 2020 and September 10, 2021, street outreach was used to recruit participants aged ≥18 or older who injected drugs within the last month and reported living in San Diego County or Tijuana, as previously described [8]. Participants were compensated $20 USD for their study visits. Protocols were approved by institutional review boards at the University of California San Diego and Xochicalco University.

### Survey Measures

After providing informed consent, participants underwent interviewer-administered surveys at baseline and approximately one week later using computer assisted personal interviews. Surveys assessed socio-demographics, substance use, chronic health conditions (e.g., diabetes, asthma, hypertension), food insecurity [13] and COVID-19 experiences, exposures, and protective behaviors (e.g., social distancing, masking).

We assessed COVID-19 misinformation and disinformation by reading a number of statements to participants and assessing the extent to which they agreed or disagreed. To assess COVID-19 misinformation, we presented participants with seven statements about SARS-CoV-2 transmission, severity, immunity, symptoms, treatments, and vaccines, and asked them to classify each statement as “True”, “False,” or “Unsure”. We then created a binary variable for each statement indicating whether the participant was misinformed or not.

We assessed COVID-19 disinformation through endorsement of six conspiracy theory items, three of which were based on work by Romer and Jamison (e.g., “COVID-19 was created by the pharmaceutical industry” or “the Chinese government”; “childhood vaccines cause autism” [7]). Based on field experience and the media, we added three new items: “COVID-19 vaccines include a tracking device”, “alter DNA”, or are being offered to communities differentially (i.e., “COVID-19 vaccines offered to ‘people like me’ are not as safe”). We dichotomized responses to indicate endorsement of disinformation

(“True” and “Unsure”) or not (“False”) and summed them into a total score ranging from 0 to 6. The mean inter-item correlation value was 0.31, which indicates optimal internal consistency [14].

Our primary outcome of interest, COVID-19 vaccine hesitancy, was assessed by asking participants whether they would agree to receive a free COVID-19 vaccine if it were offered to them. This question was introduced on 05/11/20. However, items on COVID-19 knowledge and beliefs were not introduced until 14/05/21.

### SARS-CoV-2 Antibody Detection

Serology was conducted by Genalyte® (San Diego, CA), using their Maverick™ Multi-Antigen Serology Panel [15] that detects IgG and IgM antibodies to five SARS-CoV-2 antigens (Nucleocapsid, Spike S1-S2, Spike S1, Spike S1-RBD, Spike S2) within a multiplex format based on photonic ring resonance. A machine learning algorithm was used to call results using the Random Forest Ensemble method with 3000 decision trees.[16]

### HIV and HCV Serology

Rapid HIV and HCV tests were conducted using the Miriad^®^ HIV/HCV Antibody InTec Rapid Anti-HCV Test (Avantor, Radnor, PA). Reactive and indeterminate tests underwent a second rapid test with Oraquick^®^ HIV or Oraquick^®^ HCV, respectively (Orasure, Bethlehem, PA).

### Statistical Analysis

Participants who responded that they were unsure or would not agree to receive a free COVID-19 vaccine if it were offered to them were coded as vaccine hesitant.

Characteristics of participants who were and were not COVID-19 vaccine hesitant were compared using Mann-Whitney U tests for continuous variables and Chi-square or Fisher’s Exact tests for categorical variables.

Univariate and multivariable logistic regressions with robust standard error estimation via generalized estimating equations were performed to identify factors associated with vaccine hesitancy. Variables attaining ≤10% significance were considered for inclusion in multivariable models, which is an acceptable range supported by literature [17]. In the multivariable model, only variables that retained their significance at 0.05 level were included. The multivariable model also controlled for time using a linear and quadratic term, which were highly significant but did not change the parameter estimates or their significance. We checked the final multivariable model for integrity by assessing relationships between the predictors (e.g., correlations, confounding, interactions). Multi-collinearity was assessed and ruled out by appropriate values of the largest condition index and variance inflation factors. All statistical analyses were conducted using SAS, version 9.4.

## Results

### Sample Characteristics and Vaccine Hesitancy

Of 550 participants who completed baseline and supplemental interviews, 508 (92.4%) reported that they were not vaccinated. Of these, 393 (77%) had been asked the vaccine hesitancy and COVID-19 knowledge questions and were included in this analysis. Of these 393 participants, most identified as male (74.6%) and Hispanic, Latinx, or Mexican (81.4%), and by design, approximately half (48.6%) resided in San Diego County (Table 1). Mean age was 42.4 years (standard deviation [SD]: 10.2).

**Table 1.** Characteristics Associated with COVID-19 vaccine hesitancy among PWID in San Diego, CA and Tijuana, Mexico (N=393)

| **Baseline Characteristics** | **Vaccine Hesitant**<br>**N=127** | **Not Vaccine Hesitant**<br>**N=266** | **Total**<br>**N=393** | **P** |
| --- | --- | --- | --- | --- |
| ***Socio-demographics*** |  |  |  |  |
| Male | 95(74.8%) | 198(74.4%) | 293(74.6%) | .94 |
| Mean Age (standard deviation [SD]) | 40.5(10.8) | 43.3(9.8) | 42.4(10.2) | .009 |
| Hispanic/Latinx/Mexican | 94(74.0%) | 226(85.0%) | 320(81.4%) | .009 |
| Speaks English | 91(71.7%) | 164(61.7%) | 255(64.9%) | .05 |
| Born in the US | 73(57.5%) | 89(33.5%) | 162(41.2%) | <.001 |
| Primary residence in San Diego | 73(57.5%) | 118(44.4%) | 191(48.6%) | .02 |
| Mean years of school completed (SD) | 10.1(3.0) | 9.0(3.4) | 9.3(3.3) | <.001 |
| Married or common law | 26(20.5%) | 57(21.4%) | 83(21.1%) | .83 |
| Average monthly income <500 USD | 86(67.7%) | 165(62.0%) | 251(63.9%) | .27 |
| Has a smart phone or access to a computer | 51(40.2%) | 36(13.5%) | 87(22.1%) | <.001 |
| ***Potential COVID-19 exposures*** |  |  |  |  |
| Homeless* | 65(51.2%) | 99(37.2%) | 164(41.7%) | .009 |
| Mean # of hours spent on the street (SD)* | 16.7(7.6) | 14.7(6.9) | 15.3(7.2) | .008 |
| Incarcerated* | 10(7.9%) | 20(7.5%) | 30(7.6%) | 1.00 |
| Mean # of people in the same household (SD)* | 6.2(15.2) | 5.9(11.9) | 6.0(13.0) | .04 |
| Engaged in sex work* | 15(11.8%) | 40(15.0%) | 55(14.0%) | .39 |
| Client of sex worker* | 6(4.7%) | 19(7.1%) | 25(6.4%) | .51 |
| Exposed to someone diagnosed with COVID-19 | 12(9.4%) | 12(4.5%) | 24(6.1%) | .06 |
| Income worse since COVID began | 78(61.9%) | 197(74.9%) | 275(70.7%) | .008 |
| Low/very low food security since COVID began | 98(77.2%) | 231(86.8%) | 329(83.7%) | .02 |
| ***Substance use*** |  |  |  |  |
| Smokes cigarettes | 115(90.6%) | 236(88.7%) | 351(89.3%) | .58 |
| Smoked or vaped marijuana* | 69(54.3%) | 133(50.0%) | 202(51.4%) | .42 |
| Smoked/snorted/inhaled/vaped methamphetamine* | 85(66.9%) | 139(52.3%) | 224(57.0%) | .006 |
| Smoked/snorted/inhaled crack or powder cocaine* | 20(15.7%) | 17(6.4%) | 37(9.4%) | .003 |

| **Baseline Characteristics** | **Vaccine**<br>**Hesitant**<br>**N=127** | **Not**<br>**Vaccine**<br>**Hesitant**<br>**N=266** | **Total**<br>**N=393** | **P** |
| --- | --- | --- | --- | --- |
| Smoked/snorted/inhaled/vaped either heroin or fentanyl* | 51(40.2%) | 64(24.1%) | 115(29.3%) | .001 |
| Injected methamphetamine* | 67(52.8%) | 108(40.6%) | 175(44.5%) | .02 |
| Injected cocaine* | 9(7.1%) | 17(6.4%) | 26(6.6%) | .83 |
| Injected either heroin or fentanyl* | 115(90.6%) | 241(90.6%) | 356(90.6%) | .99 |
| Mean # of years of injection drug use (SD) | 18.7(12.2) | 20.9(11.8) | 20.2(12.0) | .06 |
| Mean # of times injected drugs per day* | 2.7(1.5) | 2.4(1.5) | 2.5(1.5) | .07 |
| Visited shooting galleries* | 5(3.9%) | 32(12.0%) | 37(9.4%) | .01 |
| Receptive needle sharing* | 66(52.0%) | 150(56.4%) | 216(55.0%) | .41 |
| Crossed border to inject drugs* | 33(26.0%) | 109(41.0%) | 142(36.1%) | .004 |
| ***Health conditions*** |  |  |  |  |
| Tested HIV+ | 7(5.6%) | 26(9.8%) | 33(8.4%) | .18 |
| Tested HCV+ | 60(47.2%) | 93(35.0%) | 153(38.9%) | .02 |
| Has at least one chronic illness (excluding seasonal allergies and acne/skin problems) | 57(44.9%) | 74(27.8%) | 131(33.3%) | <.001 |
| Mean # of chronic conditions (excluding seasonal allergies and acne/skin problems; SD) | 0.9(1.5) | 0.4(0.8) | 0.6(1.1) | <.001 |
| ***Protective behaviors during the COVID-19 pandemic*** |  |  |  |  |
| Social Distancing | 41(32.3%) | 64(24.1%) | 105(26.7%) | .09 |
| Isolated or quarantined itself | 11(8.7%) | 9(3.4%) | 20(5.1%) | .05 |
| Wore face mask | 86(67.7%) | 206(77.4%) | 292(74.3%) | .04 |
| Increased handwashing/sanitizer | 29(22.8%) | 20(7.5%) | 49(12.5%) | <.001 |
| Engaged in at least one protective behavior | 105(82.7%) | 244(91.7%) | 349(88.8%) | .008 |
| ***COVID-19-related disinformation (i.e., endorsement of conspiracy theories)*** |  |  |  |  |
| Thinks that the pharmaceutical industry created the COVID-19 virus | 86(67.7%) | 117(44.0%) | 203(51.7%) | <.001 |
| Thinks that the coronavirus was created by the Chinese government as a biological weapon | 91(71.7%) | 151(56.8%) | 242(61.6%) | .005 |
| Thinks that vaccines given to children for diseases like measles and mumps cause autism | 100(78.7%) | 150(56.4%) | 250(63.6%) | <.001 |
| Thinks that COVID vaccines being offered to 'people like me' are not as safe as other COVID vaccines | 65(51.2%) | 95(35.7%) | 160(40.7%) | .004 |
| Thinks that COVID vaccines include a tracking device | 66(52.0%) | 69(25.9%) | 135(34.4%) | <.001 |
| Thinks that some COVID vaccines could change their DNA | 57(44.9%) | 64(24.1%) | 121(30.8%) | <.001 |
| Mean # of conspiracies they believe (out of 6) (SD) | 3.7(1.6) | 2.4(1.9) | 2.8(1.9) | <.001 |
| ***COVID-19-related misinformation (i.e., incorrect knowledge items)*** |  |  |  |  |
| Does not think that the virus that causes COVID-19 can be easily spread from one person to another | 30(23.6%) | 54(20.3%) | 84(21.4%) | .45 |
| Does not think that many thousands of people have died from COVID-19 | 19(15.0%) | 23(8.6%) | 42(10.7%) | .06 |
| Thinks that most people already have immunity to COVID-19 | 85(66.9%) | 165(62.0%) | 250(63.6%) | .35 |
| Thinks that you can tell someone has COVID-19 by looking at them | 48(37.8%) | 66(24.8%) | 114(29.0%) | .008 |
| Thinks that there are effective treatments for COVID-19 that can cure most people | 96(75.6%) | 211(79.3%) | 307(78.1%) | .40 |
| Thinks that having COVID-19 is about as dangerous as having the flu | 82(64.6%) | 169(63.5%) | 251(63.9%) | .84 |
| Does not think that COVID vaccines are safe for pregnant women | 109(85.8%) | 141(53.0%) | 250(63.6%) | <.001 |
| ***Most important source of COVID-19-related information*** |  |  |  |  |
| Friends<sup>Y1</sup> | 59(48.0%) | 137(52.1%) | 196(50.8%) | .45 |
| Doctors/health professionals<sup>Y1</sup> | 12(9.8%) | 9(3.4%) | 21(5.4%) | .01 |
| Social media<sup>Y1</sup> | 26(21.1%) | 21(8.0%) | 47(12.2%) | <.001 |
| ***Additional COVID-19-related experiences*** |  |  |  |  |
| Knows someone who died from covid-19 | 45(35.4%) | 77(28.9%) | 122(31.0%) | .19 |
| Mean for: On a scale of 1 (low) to 10 (very), how worried are you of getting COVID-19 (or getting it again; SD) | 4.0(3.2) | 5.4(2.9) | 5.0(3.1) | <.001 |
| Knows someone who has been vaccinated for COVID-19 | 80(63.0%) | 160(60.2%) | 240(61.1%) | .59 |
| Thinks they had COVID-19 | 21(16.5%) | 16(6.0%) | 37(9.4%) | <.001 |
| Has been tested for COVID-19 test outside of our study | 40(31.5%) | 38(14.3%) | 78(19.8%) | <.001 |
| Has been exposed to somebody with a positive COVID-19 test result | 12(9.4%) | 12(4.5%) | 24(6.1%) | .06 |
| Had at least one COVID-19 symptom on day of interview | 39(30.7%) | 69(25.9%) | 108(27.5%) | .32 |
| Tested SARS-CoV-2 seropositive <sup>Y2</sup> | 48(42.9%) | 97(39.8%) | 145(40.7%) | .58 |
| Ever had a flu vaccine | 59(46.8%) | 118(44.9%) | 177(45.5%) | .72 |

\*Past 6 months; Missing values: <sup>Y1</sup> n=6, <sup>Y2</sup> n=33;

In the past six months, most participants (90.6%) injected either heroin or fentanyl, and 40.6% injected methamphetamine. Additional non-injection substance use involved smoking cigarettes (89.3%), methamphetamine (57.0%), marijuana (51.4%) and heroin or fentanyl (29.3%). Over one third were HCV-seropositive (38.9%), 8.4% were HIV-seropositive, and 33.3% reported at least one other chronic health condition.

Most participants reported reduced food security (83.7%) and income (70.7%) since the COVID-19 pandemic began, and 40.7% tested SARS-CoV-2 seropositive. Substantial proportions of participants reported knowing someone who had died from COVID-19 (31.0%) and knowing someone who had been vaccinated for COVID-19 (61.1%). Overall, 22.1% had access to a smart phone (19.3%) or computer (9.7%).

Overall, about one third of participants were vaccine hesitant (n=127, 32.3%). Compared to those who were willing to be vaccinated against COVID-19 (Table 2), vaccine-hesitant participants were younger (mean age: 40.5 vs. 43.3 years, *p*=.009) and had higher education (mean years of schooling completed: 10.1 vs. 9.0, *p*<.001). Higher proportions of vaccine-hesitant participants were born in the U.S. (57.5% vs. 33.5%, *p*<.001), currently resided in San Diego (57.5% vs. 44.4%, *p*=.01), and were homeless (51.2% vs. 37.2%, *p*=.009). Compared to other participants, those who felt that they had already had COVID-19 were more likely to be vaccine-hesitant (16.5% vs. 6%, p<0.001).

**Table 2:** Factors associated with SARS-CoV-2 vaccine hesitancy in Tijuana and San Diego.

| **Baseline characteristics** | **Univariate OR (95% CI)** |
| --- | --- |
| ***Socio-demographics*** |  |
| Male | 1.02 ( 0.63,1.66 ) |
| Age<sup>P</sup> | 0.97 ( 0.95,1.00 ) |
| Hispanic/Latinx/Mexican<sup>P</sup> | 0.50 ( 0.30,0.85 ) |
| Speaks English<sup>P</sup> | 1.57 ( 0.99,2.49 ) |
| Born in the US<sup>P</sup> | 2.69 ( 1.74,4.15 ) |
| Primary residence in San Diego | 1.70 ( 1.11,2.60 ) |
| Highest year of school completed<sup>¥P</sup> | 1.12 ( 1.04,1.20 ) |
| Married or common law | 0.94 ( 0.56,1.59 ) |
| Monthly income <500 USD | 1.28 ( 0.82,2.01 ) |
| Has a smart phone or access to a computer<sup>P</sup> | 4.29 ( 2.60,7.06 ) |
| ***Potential COVID-19 exposures*** |  |
| Homeless*<sup>P</sup> | 1.77 ( 1.15,2.71 ) |
| # of hours spent on the street*<sup>P</sup> | 1.04 ( 1.01,1.08 ) |
| Incarcerated* | 1.05 ( 0.48,2.32 ) |
| # of people in the same household*<sup>¥P</sup> | 1.00 ( 0.99,1.02 ) |
| Engaged in sex work* | 0.76 ( 0.40,1.43 ) |
| Client of sex worker* | 0.64 ( 0.25,1.66 ) |
| Exposed to someone diagnosed with COVID-19<sup>P</sup> | 2.21 ( 0.96,5.06 ) |
| Income worse since COVID began<sup>P</sup> | 0.54 ( 0.35,0.86 ) |
| Low or very low food security since COVID began<sup>P</sup> | 0.51 ( 0.30,0.88 ) |
| ***Substance use*** |  |
| Smokes cigarettes | 1.22 ( 0.60,2.47 ) |
| Smoked or vaped marijuana*<sup>P</sup> | 1.19 ( 0.78,1.82 ) |
| Smoked/snorted/inhaled/vaped methamphetamine*<sup>P</sup> | 1.85 ( 1.19,2.87 ) |
| Smoked/snorted/inhaled crack or powder cocaine*<sup>P</sup> | 2.74 ( 1.38,5.43 ) |
| Smoked/snorted/inhaled/vaped heroin or fentanyl*<sup>P</sup> | 2.12 ( 1.35,3.33 ) |
| Injected methamphetamine* | 1.63 ( 1.07,2.50 ) |
| Injected cocaine* | 1.12 ( 0.48,2.58 ) |
| Injected heroin or fentanyl* | 0.99 ( 0.48,2.05 ) |
| Years of injection drug use<sup>yp</sup> | 0.98 ( 0.97,1.00 ) |
| #Times injected drugs per day<sup>yp</sup> | 1.12 ( 0.97,1.29 ) |
| Visited shooting galleries*<sup>p</sup> | 0.30 ( 0.11,0.79 ) |
| Receptive needle sharing* | 0.84 ( 0.55,1.28 ) |
| Crossed border to inject drugs*<sup>p</sup> | 0.51 ( 0.32,0.81 ) |
| ***Health conditions*** |  |
| Tested HIV+ | 0.54 ( 0.23,1.29 ) |
| Tested HCV+<sup>p</sup> | 1.67 ( 1.08,2.56 ) |
| Has at least one chronic condition (excluding seasonal allergies and acne/skin problems)<sup>p</sup> | 2.11 ( 1.36,3.28 ) |
| # of chronic conditions (excluding seasonal allergies and acne/skin problems)<sup>p</sup> | 1.49 ( 1.23,1.80 ) |
| ***Protective behaviors during the COVID-19 pandemic*** |  |
| Practiced Social distancing<sup>p</sup> | 1.50 ( 0.94,2.40 ) |
| Isolated or quarantined itself<sup>p</sup> | 2.71 ( 1.09,6.71 ) |
| Wore face mask<sup>p</sup> | 0.61 ( 0.38,0.98 ) |
| Increased handwashing/sanitizer<sup>p</sup> | 3.64 ( 1.97,6.74 ) |
| Engaged in at least one protective behavior<sup>p</sup> | 0.43 ( 0.23,0.81 ) |
| ***COVID-19-related disinformation (i.e., endorsement of conspiracy theories)*** |  |
| Thinks that the pharmaceutical industry created the COVID-19 virus<sup>p</sup> | 2.67 ( 1.71,4.16 ) |
| Thinks that the coronavirus was created by the Chinese government as a biological weapon<sup>p</sup> | 1.93 ( 1.22,3.04 ) |
| Thinks that vaccines given to children for diseases like measles and mumps cause autism<sup>p</sup> | 2.86 ( 1.76,4.67 ) |
| Thinks that COVID vaccines being offered to people like me are not as safe as other COVID vaccines<sup>p</sup> | 1.89 ( 1.23,2.90 ) |
| Thinks that COVID vaccines include a tracking device<sup>p</sup> | 3.09 ( 1.98,4.81 ) |
| Thinks that some COVID vaccines could change their DNA<sup>p</sup> | 2.57 ( 1.64,4.03 ) |
| # of conspiracy items that they believe (out of six)<sup>p</sup> | 1.44 ( 1.28,1.62 ) |
| ***COVID-19-related misinformation (i.e., incorrect knowledge items)*** |  |
| Does not think the virus that causes COVID-19 can be easily spread from one person to another | 1.21 ( 0.73,2.02 ) |
| Does not think that many thousands of people have died from COVID-19<sup>P</sup> | 1.86 ( 0.97,3.56 ) |
| Thinks that most people already have immunity to COVID-19 | 1.24 ( 0.79,1.93 ) |
| Thinks that you can tell someone has COVID-19 by looking at them<sup>P</sup> | 1.84 ( 1.17,2.90 ) |
| Thinks that there are effective treatments for COVID-19 that can cure most people<sup>P</sup> | 0.81 ( 0.49,1.33 ) |
| Thinks that having COVID-19 is about as dangerous as having the flu | 1.05 ( 0.67,1.63 ) |
| Does not think that COVID vaccines are safe for pregnant women<sup>P</sup> | 5.37 ( 3.09,9.34 ) |
| ***Most important source of COVID-19-related information*** |  |
| Friends<sup>Y1</sup> | 0.85 ( 0.55,1.30 ) |
| Doctors/health professionals<sup>Y1P</sup> | 3.05 ( 1.25,7.45 ) |
| Social media<sup>Y1P</sup> | 3.09 ( 1.66,5.75 ) |
| ***Additional COVID-19-related experiences*** |  |
| Knows someone who died of COVID-19 | 1.35 ( 0.86,2.11 ) |
| On a scale of 1 (low) to 10 (very), how worried are you of getting COVID-19 (or getting it again)<sup>P</sup> | 0.86 ( 0.79,0.93 ) |
| Knows someone who has been vaccinated for COVID-19 | 1.13 ( 0.73,1.74 ) |
| Thinks they had COVID-19<sup>P</sup> | 3.10 ( 1.55,6.16 ) |
| Has been tested for COVID-19 outside of our study<sup>P</sup> | 2.76 ( 1.66,4.59 ) |
| Has been exposed to somebody with a positive COVID-19 test result<sup>P</sup> | 2.21 ( 0.96,5.06 ) |
| Had at least one COVID-19 symptom day of interview | 1.27 ( 0.79,2.02 ) |
| Tested SARS-CoV-2 seropositive<sup>Y2</sup> | 1.14 ( 0.72,1.79 ) |
| Ever had a flu vaccine | 1.08 ( 0.71,1.66 ) |

\*past 6 months; Missing values: <sup>Y1</sup> n=6, <sup>Y2</sup> n=33; <sup>Y</sup>Per one unit increase; <sup>P</sup>P-value<0.10

Almost all participants endorsed at least one statement that reflected COVID-19 misinformation (99%), such as thinking that COVID-19 is about as dangerous as having the flu (63.9%). Respondents who thought COVID-19 vaccines were unsafe for pregnant women or believed they could tell if someone had COVID-19 by looking at them were significantly more likely to be vaccine hesitant.

A majority of participants also endorsed at least one conspiracy theory related to COVID-19 or vaccines (85%). COVID-19 disinformation scores were higher among those who were vaccine hesitant (mean number of COVID-19 conspiracy theories endorsed out of six total: 3.7 vs. 2.4, *p*<.001). Vaccine-hesitant participants were more likely to identify social media as their primary source of COVID-19-related information (21.1% vs. 8%, *p*<.001). There were no differences observed in vaccine hesitancy related to identifying friends as a primary source of COVID-19 information (50.8% overall) or ever having had a flu vaccine (45.5% overall).

### COVID-19-Related Correlates of Vaccine Hesitancy

Participants who engaged in at least one protective behavior (e.g., social distancing, isolating oneself, wearing masks, increasing handwashing), were significantly less likely to be vaccine hesitant (unadjusted odds ratio [OR]: 0.43; 95% confidence interval [CI]: 0.23-0.81; Table 2). Those who were more worried about getting COVID-19 were also less likely to be vaccine hesitant (OR: 0.86 per point increase; 95% CI: 0.79-0.93). Conversely, those who thought they had had COVID-19 and had been tested for COVID-19 outside of this study were significantly more likely to be vaccine hesitant, and having been exposed to somebody testing positive for COVID-19 was marginally associated with higher unadjusted odds of vaccine hesitancy (OR: 2.21; CI: 0.96-5.06; *p*=.06). Regarding other COVID-19-related experiences and exposures, having COVID-19 symptoms on the day of the interview or knowing someone else who had been vaccinated against COVID-19 were not associated with vaccine hesitancy. Testing SARS-CoV-2 seropositive in our study was not associated with vaccine hesitancy although participants were unaware of their test results at the time of interview.

### Factors Independently Associated with Vaccine Hesitancy

In our final multivariate model controlling for time (Table 3), older age was inversely associated with vaccine hesitancy (adjusted OR [aOR]: 0.97 per year increase in age; CI: 0.95-0.99). Participants with greater food insecurity and concern about acquiring SARS-CoV-2 were less likely to be vaccine hesitant (aOR: 0.44; CI: 0.23-0.87; and aOR: 0.85 per point increase; CI: 0.77-0.93, respectively). Greater numbers of chronic health conditions were independently associated with vaccine hesitancy (aOR: 1.46 per additional chronic condition; CI: 1.17-1.82). Those with a smart phone or computer access were almost four times more likely to be vaccine hesitant (aOR: 3.75; CI: 2.07-6.82). Citing social media as one’s most important source of COVID-19 information was marginally associated with vaccine hesitancy (aOR: 1.86; CI: 0.94-3.70, p=0.07). Finally, COVID-19-related disinformation was independently associated with vaccine hesitancy (aOR: 1.51 per additional conspiracy theory endorsed; CI: 1.31-1.74).

**Table 3:** Factors Independently Associated with COVID-19 Vaccine Hesitancy among People who Inject Drugs in San Diego, CA and Tijuana, Mexico.

| **Baseline characteristics** | **Adjusted OR\*\***<br>**(95% CI)** |
| --- | --- |
| Age<sup>¥</sup> | 0.97 ( 0.95,1.00 ) |
| Has low/very low food security | 0.44 ( 0.23,0.87 ) |
| Has a smart phone or access to a computer | 3.75 ( 2.07,6.82 ) |
| The number of self-reported chronic conditions (excluding allergies and acne/other skin conditions)<sup>¥</sup> | 1.46 ( 1.17,1.82 ) |
| Number of conspiracies they believe (out of 6)<sup>¥</sup> | 1.51 ( 1.31,1.74 ) |
| Most important source of COVID-19 information: social media<sup>¥1</sup> | 1.86 ( 0.93,3.70 ) |
| On a scale of 1 to 10, how worried are you of getting COVID-19 (or getting it again)<sup>¥</sup> | 0.85 ( 0.77,0.93 ) |

\*\*variables in the multivariable model were adjusted for all the variables in the model as well as for time; Missing values:<sup>¥1</sup> n=6; ¥Per one unit increase;

In a sub-analysis that included the 42 participants who reported having had at least one COVID-19 vaccine dose and coded them as willing, parameter estimates in our final model were essentially unchanged, with the exception of primarily obtaining COVID-19 information from social media, which became highly significant.

## Discussion

Vaccine hesitancy is a critical challenge to COVID-19 pandemic control efforts, especially for vulnerable populations including people who use and inject drugs. In our community-based sample of PWID in the San Diego-Tijuana border region, nearly one third of participants were hesitant about COVID-19 vaccines and almost all endorsed statements reflecting COVID-19 misinformation or disinformation. While the dissemination of COVID-19 disinformation on social media has been reported as undermining vaccine uptake in the general population [3], we found that it is also influential in a disadvantaged population that has limited access to the Internet. Our analysis also identified specific intervention targets that provide avenues for improving vaccine trust and uptake in this socially marginalized population.

We found that COVID-19 disinformation, operationalized as endorsement of specific COVID-19 related conspiracy theories, was independently associated with vaccine hesitancy, while COVID-19 misinformation was not. While research on COVID-19 vaccination hesitancy among substance using populations remain scarce, a study conducted with substance use disorder treatment patients also failed to link COVID-19 knowledge with trust in vaccines [18]. These findings imply that disinformation may be a stronger driver of COVID-19 vaccine hesitancy than misinformation among PWID.

The significant role of COVID-19 disinformation in influencing vaccine hesitancy extends a long-established foundation of medical mistrust among PWID who often avoid traditional clinical settings, preferring to receive prevention information and services in community-based settings [11]. This distrust, along with preferences for alternative sources of information, may lead some individuals to seek health advice online or through social media, where more false information may exist than factual, evidence-based information [19], and where COVID-19-related disinformation has been perpetuated [3].

Having access to smart phones or computers was strongly associated with COVID-19 vaccine hesitancy in our sample, despite the fact that only 22% had access to either. PWID in other community-based studies increasingly report having regular access to mobile phones and the Internet, particularly within public spaces [12, 20, 21]. In our study, citing social media as one’s most important source of COVID-19 information was marginally associated with vaccine hesitancy, even after controlling for phone/computer access. When we repeated our analysis to include participants who had received at least one COVID-19 vaccine dose by September 10, 2021, the association between citing social media as one’s most important source of COVID-19 information and vaccine hesitancy was even stronger.

Concerns about vaccine safety were also apparent in our sample, confirming previous research that PWID, like a sizable segment of the general population [22], harbor concerns that COVID-19 vaccines are unsafe and have been tested insufficiently [23]. Taken together with our findings regarding the independent influences of disinformation and social media on vaccine hesitancy, these concerns suggest that vaccine-related educational interventions, whether delivered online or in-person (particularly for PWID with poor access social media or the Internet), should build knowledge of vaccine development processes, address concerns about medication interactions and side-effects, and increase personalized knowledge of and perceived risk of COVID-19 [24].

Increasing confidence in vaccine safety and efficacy will be particularly important for subgroups of PWID with higher levels of skepticism or susceptibility to confirmation bias (i.e., the tendency to believe information that aligns with one’s existing beliefs or experiences) [25]. As others have argued [26], interventions will be most acceptable to PWID if they are delivered by trusted sources of health information and support, such as harm reduction outreach workers, street medicine providers, recovery coaches, peers, or staff of other community-based organizations that are frequented and trusted by this population, like shelters, community centers, hostels, libraries, and other public spaces.

Interestingly, we found no evidence to support our hypothesis that Latinx participants were more likely to be vaccine hesitant, as has been reported in the literature [2, 7]. In fact, our unadjusted analysis found that PWID who were White and those born or living in San Diego were more likely to be vaccine hesitant than those who were Latinx, or born or living in Mexico. Contrary to general population-based samples in the United States [27], we did not find educational attainment to be independently associated with vaccine hesitancy.

PWID reporting food insecurity and those with higher levels of concern about SARS-CoV-2 were less hesitant about COVID-19 vaccines. However, two thirds of our sample reported willingness to get vaccinated, but only 7.6% had received at least one dose by September 10, 2021, highlighting the need to increase access. In addition, participants who had a greater number of comorbidities such as diabetes and hypertension were more vaccine hesitant than others. This is concerning since these participants are more likely to suffer severe complications from SARS-CoV-2 infection and are precisely those most in need of protection.

Our findings suggest that structural supports including financial incentives for vaccination that have been successful with other vulnerable populations could also be beneficial for PWID [28]. These include transportation assistance, co-location with other routinely accessed services (e.g., syringe exchanges, food banks, soup kitchens), concurrent vaccination of peers and family members, and vaccine administration by an expanded group of healthcare and lay providers (e.g., emergency department and drug treatment clinic personnel, community health workers). Modest financial incentives via conditional cash transfers and contingency management have demonstrated success in increasing adherence to three-dose hepatitis B vaccines among PWID and other substance using populations [29, 30].

Limitations of this study include the cross-sectional nature of the analysis, which precludes our ability to determine causal associations. Although this was a binational study, sampling was non-random and results may not generalize to other samples of PWID. We also relied on self-report and recall for many behaviors, which may have been subject to socially desirable responding. Although the COVID-19 disinformation scale we utilized had good internal consistency, other COVID-19 related knowledge measures have only been recently developed and, to our knowledge, have not been validated in this or other populations impacted by substance use. Our analysis excluded participants who were recruited before survey items on COVID-19 knowledge and vaccine hesitancy were developed. Since attitudes to COVID-19 vaccines may have changed over time, we controlled for time in our analysis. Future longitudinal, qualitative, and intervention-development studies are needed to better understand contextual factors influencing vaccine hesitancy in this population to identify strategies to best address these intervention targets.

In conclusion, we identified a concerning level of COVID-19 vaccine hesitancy among community-recruited PWID in the San Diego-Tijuana border region, which was associated with COVID-19 related disinformation, reliance on social media as a source health information, younger age and co-morbidities. Interventions that increase accurate COVID-19 vaccine knowledge, trust and motivation, while also reducing structural barriers to vaccine access are urgently needed to reduce morbidity and mortality from SARS-CoV-2 infection in this vulnerable population.

## Data Availability

De-identified data used in this analysis is available after completion of the study in May 2022. Interested parties should contact Daniela Abramovitz at for more information on how to submit a data request.

## Funding

This work was supported by the National Institute on Drug Abuse (NIDA) at the National Institutes of Health (NIH) (R01DA049644-S1, R01DA049644-02S2, K01DA043412). Additional support was provided by the National Institute of Allergy and Infectious Diseases at NIH (P30 AI036214) and by the California HIV/AIDS Research Program (CHRP) (OS17-SD-001).

## Acknowledgements

The authors gratefully acknowledge the La Frontera study team and participants in San Diego and Tijuana and staff at Genalyte and Fluxergy for assistance interpreting laboratory results, laboratory staff at the Center for AIDS Research and Sharon Park for assistance with manuscript preparation.

## Author Contributions

SAS designed the study and most survey instruments, conceived of the research questions wrote and edited the manuscript. DA conducted the data analysis, prepared the results and edited the manuscript. GR helped design the study, oversaw data collection in Tijuana and edited the manuscript. AHV designed and pre-tested the survey, oversaw collection of laboratory specimens and edited the manuscript. CV oversaw and participated in data collection in San Diego and edited the manuscript. IA programmed the study instrument, oversaw data management and edited the manuscript. TP helped design the study and survey instruments and edited the manuscript. RM assisted with the literature review and edited the manuscript. ARB helped interpret the analysis and wrote and edited the manuscript.

## Declaration of Interests

The authors report no conflicts of interest.

## Notes

### Competing Interest Statement

The authors have declared no competing interest.

### Author Declarations

IRB of University of California San Diego and the IRB of Universidad Xochicalco gave ethical approval for this work.

